# Dying to make a care policy: Community perception of causes of deaths using standardized verbal autopsy method: Saudi type 2 diabetes mellitus register data

**DOI:** 10.1101/2023.02.16.23286020

**Authors:** Faleh Alyazidi, Deler Shakely, Max Petzold, Fawaz Alyazidi, Laith Hussain-Alkhateeb

**Affiliations:** School of Public Health and Community Medicine, Sahlgrenska Academy, University of Gothenburg, Gothenburg, Sweden; Department of Public Health, College of Health Sciences at Al-Leith, Umm Al-Qura University, Al-Leith, Kingdom of Saudi Arabia; Infectious Diseases Control Department, Executive Directorate of Preventive Medicine, Makkah Healthcare Cluster, Makkah, Kingdom of Saudi Arabia

## Abstract

Diabetes mellitus (DM) is a serious global health issue which significantly impacts public health and socioeconomic development. Exploring how the community perceives the causes of deaths and their associated risk factors is crucial for public health. This study combines verbal autopsy (VA) with the Type 2 Diabetes Mellitus (T2DM) register to explore community perceptions of causes of death and associated influential factors in the western region of Saudi Arabia. 302 VA interviews were conducted with relatives or caregivers of deceased who died between 2017 and 2021 based on T2DM medical register from Alnoor Specialist Hospital in Makkah city, Saudi Arabia. Cause-specific mortality fractions (CSMFs) obtained from the VA using the InterVA-5 model were utilized to assess community perception. We used a multivariable logistic regression model to determine factors influencing community perceptions of causes of death. Lin’s CCC with 95% CI was used to analyze the concordance for the CSMFs from verbal autopsy causes of death (VACoD) as a presumed reference standard and family-reported causes of death (FRCoD). The outcomes of this study demonstrate a generally broad spectrum of community perceived mortalities, with some critical misconceptions based on the type of death and the deceased’s background and characteristics, with an overall CCC of 0.60 (95% CI: 0.20-1.00; p=003). The study findings demonstrate that community perception is weak if the deceased was male compared to female (aOR: 0.52; 95% CI: 0.26 –1.03) and if the deceased was >=80 years compared to 34–59 years (aOR: 0.48; 95% CI: 0.16-1.38), but it significantly improves among married compared to single (aOR: 2.13; 95% CI: 1.02 –4.42). The Saudi community perception of causes of death with reported type 2 diabetes was relatively plausible but varied substantially based on the type of death, sex, age >=80 years, and other vital events like marital status. Higher or lower community perception is attributed to how people may perceive risk factors associated with the causes of death, which can guide public health planning and interventional programs. The study findings further emphasize the need to employ robust and standardized VA methods within the routine medical services for a systemized assessment of families’ reported causes of death.

## Introduction

Diabetes mellitus (DM) remains a major global public health issue with a significant impact on public health and socio-economic development. Diabetes is one of the top ten causes of death worldwide. Together with cardiovascular diseases, respiratory diseases and cancer, these conditions account for over 80% of all premature non-communicable diseases (NCDs) deaths [1]. The International Diabetes Federation estimates that 536.6 million people were living with diabetes in 2021, and this number is projected to increase by 46%, reaching 783.2 million in 2045 [2], with approximately 50% of all diabetic individuals are unaware of their conditions [3]. Several reports have attributed the prevalence of DM in the Middle East and North Africa regions to the increased per capita income, urbanization, and pronounced lifestyle changes that enabled physical inactivity and increased the obesity rate.

In the context of chronic illnesses and treatment management, community perception of health and illness – which is the ability to perceive the world through sensory organs [4] – is critical for public health administrative practices. A comprehensive and integrated approach to understanding how communities perceive health and disease is crucial for developing relevant and effective health intervention strategies and can empower individuals to seek and handle health knowledge properly [5-7]. Individuals are more likely to take action to prevent illnesses if they acknowledge their susceptibility to a particular form of illness or if they are likely to experience serious consequences [8]. Lay concepts of health and illness provide essential information to biomedical models [9, 10] and can significantly impact communities’ health and illness behaviors, which requires systematic means of assessment at population level [6, 10]. Saudi Arabia is associated with this global epidemic of NCDs [11, 12]. According to the World Health Organization (WHO), Saudi Arabia has the second highest rate of diabetes in the Middle East (7^th^ highest in the world), with an estimated population of 7 million diabetics and more than 3 million pre-diabetics [13], which is a serious public health issue. Previous Saudi reports have shown that the success of national response strategies is heavily dependent on the public’s perceptions and attitudes toward the risk of an epidemic and the importance of preventive measures [14]. Addressing the health literacy problems among a culturally diverged Saudi population will be crucial by virtue of the undergoing health system re-engineering towards the Saudi 2030 Vision. The Vision 2030 Health Sector Transformation Program was initiated in 2021 with a five-year goal of reforming the health sector to become a comprehensive, effective, and integrated health system that prioritizes the health of individual and society, including citizens, residents, and visitors [15]. A growing body of evidence have additionally demonstrated that inadequate health literacy is significant among the Saudi population [16-18], whereby negative perceptions of diabetes risk factors can impede disease management and prevention. Patients with poor health literacy skills are prevalent among the Saudi population. A published cross-sectional survey conducted in Saudi Arabia to describe the distribution of low health literacy in the Saudi population found that 46% of respondents were classified as having low health literacy; this was associated with older age groups [17]. This group demands actively engaged healthcare professionals to effectively simplify the care management, which will only add more burden to the already overstretched health system caused by COVID-19 and other emerging diseases.

Global statistics show that about half of annual deaths pass without formal medical certification leaving crucial causes of death information outside any form of useful application for policy making and public health program monitoring [19-23]. While this issue is broadly associated with low- and middle-income countries (LMICs) due to insufficient investment in civil registration and vital statistics (CRVS) systems [23], higher income countries are also prone to misclassification of crucial international classification of disease (ICD) information [24]. Saudi Arabia is among countries facing demographic transition, which can influence the doctor-patient communications and hence the ultimate uptake of health information and their classifications.

Verbal autopsy (VA) – standardized interviews conducted by trained fieldworkers or healthcare staff with final caregivers or close relatives of the deceased on the medical signs, symptoms, and circumstances surrounding deaths – has emerged in more recent time as a widely used method for determining causes of death in settings where adequate reporting and reliable classification of causes of death are distracted [25, 26]. While this set of VA information is classically processed by physicians to determine a likely medical cause of death, computerized tools such as *InterVA-5* have been validated in several settings, which demonstrate significant advantages over the VA physician review process, mainly being cheap and consistent over time and place [6, 27, 28]. VA is commonly conducted within registered and unregistered population and can consistently elicit information through structured and narrative sections using the standardized WHO VA questionnaires. At the end of the VA interview, the interviewer would typically finish off with asking respondents about their own interpretation of the causes of death of their deceased, such information that can form the basis of understanding community perception of causes of death. Social autopsy (SA), a less structured method for investigating death, includes questions on changeable social, cultural, and healthcare system aspects, which shows great potential for routine public health applications [23, 29]. SA has often been used in connection with VA in registered populations. Nevertheless, it tends to be time and cost consuming if used as a sole tool, and unlike VA, it will not be amenable for large-scale applications or automated interpretations, which led to its innovation as VA-SA integrated process [29, 30].

Within the Saudi research and health policy interests, where VA is rarely practiced, no studies have yet sought standardized tools such as the VA to explore the community perception of causes of death, which can have significant implications in guiding public health planning and novel interventional programs. This study will employ VA to explore the community perceptions of causes of death based on T2DM register in the Western region of Saudi Arabia. To success this study aim, VA causes of death (VACoD) as a presumed reference standard will be compared to the family-reported causes of death (FRCoD), and the determinants of the community perception will be further assessed across different causes of death groups and population backgrounds and characteristics.

## Methodology

### Study population and context

The study conducted 302 Verbal autopsy interviews with relatives or caregivers of the deceased who died between 2017 and 2021, according to T2DM medical register from the Alnoor Specialist Hospital – a public hospital in Makkah city, Saudi Arabia, which is among three other hospitals that constitute to major healthcare provider to a population size of 2.5 million. All patients included in this study fulfilled the study criteria; (1) death occurred between 2017 and 2021 (small history interval to reduce recall bias during the VA process), (2) at least one clinical diagnosis (ICD) of T2DM, and (3) informed consent to perform the verbal autopsy interview given by the respondent.

Makkah is a large province and of a relatively dynamic population compared to other Saudi provinces, mainly due to their close and constant interaction with people who come from all over the world to perform religion pilgrimage (Hajj and Umrah). This frequent (short and long-term) stay of a large population-scale creates a special demographic mix where migration of culture, economy, diseases, and demography is highly prevalent in this region. This diverged community has the potential to influence the community perceptions and beliefs about a certain phenomenon, such as how they perceive illness or causes of death.

### Verbal autopsy interpretation

For VA interpretation, several software programs are used for automated interpretation such as *‘Smart VA’, ‘InSilicoVA’*, and *‘InterVA’* [31-33], with the *InterVA* being used more broadly [19, 33]. The 2016 WHO VA instrument has been updated to contain all the input variables needed by all three software, making their use more consistent and standardized. [28, 34, 35]. In recent years, a series of InterVA models have been created to interpret VA data using Bayesian probabilistic modeling [36].

VA uses a Bayesian probability model to determine the most probable cause of death and their corresponding likelihoods for each VA case [5, 26, 37]. The Bayesian concept operation permits the combination of medical experts’ views as prior and relevant available data, which can collectively augment the model outputs [6, 38]. This methodological process used in the InterVA model is explained in full elsewhere [36]. Using Bayes theorem, up to three probable causes of death (likelihood) are typically assigned for each cause by the automated InterVA tool, given the specific signs and symptoms reported [26, 33]. Each assigned cause has corresponding likelihoods, and the sum likelihoods of all assigned causes has a maximum value of 1.00 (or 100%). In cases where the likelihood proportions did not add up to 1.0 for a particular case, any remaining margin of likelihood not accounted for by the likelihood of the first, second, and third causes can therefore be included as a partial indeterminate component in analyzing overall cause of death and CSMFs patterns. It has been suggested that this is a more practical approach than aggregating the sum of small residual probabilities of unlikely causes, which might lead to misleading conclusion in a large number of cases [36]. The Cause-Specific Mortality Fraction (CSMF) is an output from the algorithm. CSMF corresponds to the proportion that each cause of death category contributed to the total number of deaths [5, 6, 38].

### Data collection management

Information on the deceased’s background and characteristics were taken directly from the medical register at Alnoor Specialist hospital. Death records contained data for capturing vital events [deaths in and out-hospital, date of death, sex, age, ICD-10 code of causes of death, and contact details of the next of kin]. Considering the heterogeneous and dynamic population in this Province, other characteristics of the deceased such as nationality, marital status, and education, were also considered. A total of pre-estimated sample of 302 relatives or close caregivers of the deceased were randomly chosen from the medical register and consented to conduct the VA interview. In addition, interviews were administered by the first author together with a trained research assistant. The standard face-to-face VA interview process has been challenged by the COVID-19 pandemic, which coincided with the strict physical distancing preventive measures implemented in Saudi Arabia. Therefore, telephone interviews were sought as a substitute for the standard face-to-face method. The interviews were conducted in Arabic without audio-recording, but data were reviewed and safely stored at the university electronic databases after removing all personal identifiers, and all participants were anonymized. Closed-ended questions were collected using the Open Data Kit (ODK) on Android smartphones. VA data were reported into a Microsoft Access database, and the participants’ study IDs were used to link the VA data with the background and characteristic information.

### Data management

The deceased’s age was categorized into four age groups (34-59, 60-69, 70-79, and >=80 years old), and the levels of education were grouped as *illiterate, basic education including (primary, secondary school, and high school), and advanced education including* university level. The relationship between the VA informants and the deceased was classified into two groups (first-degree relative and second-degree relative), whereby the deceased marital status was dichotomized as single and married. The place of death was recorded and processed as binary information (home, hospital/health facility). The causes of death were further categorized into broader categories, described in S1 Table.

### Data analysis

Numerical and graphical descriptive statistics were generally sought for causes of death data in overall and per groups. Each individual VA was processed using InterVA-5 model, which assigns up to three likely causes of death per WHO VA cause categories [39], while FRCoDs were given as a single or two causes of death. Crude and adjusted logistic regression for the binomial agreement (agreement or not) between VACoD and FRCoD was analyzed against the informant’s background, characteristics, and socioeconomic factors for each cause of death category. All potential confounders from the background and characteristic factors were assessed in a forward-wise likelihood ratio test. Statistical analyses were performed using STATA release 17.0 software using a significant level of 5% as a cut-off for the hypothesis test.

Generated causes of death likelihoods were utilized to calculate population-level CSMFs for each cause of death group between VACoD and FRCoD, and their proportional absolute differences and 95% confidence intervals were calculated. A concordance correlation coefficient (CCC) for the CSMFs from VACoD and FRCoD was also calculated using Lin’s CCC with 95% CI [40]. Based on Pearson’s correlation, the CCC compares the viewpoints between the two measures – just like other correlation tests, CCC has a range of -1 to 1, with 1 representing perfect agreement [41]. The CCC combines precision and accuracy measures to determine how far the observed data vary from the perfect concordance line (that is, the line at 45 degrees on a square scatter plot). Lin’s coefficient increases in value as a function of the data’s reduced major axis’s closeness to the perfect concordance line (data accuracy) and the data’s tightness about its reduced major axis (the precision of the data) [40]. The CCC can essentially estimate the agreement between VACOD and FRCOD from a population causes of death level.

### Ethical approval

This study received the ethical approval to perform the VA interviews from the Institutional Review Board (IRB) at Makkah Health Affairs, Saudi Arabia (IRB Number: H-02-K-076-0321-478). The study was aligned with the Declaration of Helsinki. Informed consent was fully respected at both individual and household levels, with the right to refuse or withdraw from interviews. Additionally, data was only accessed by the principal researcher for the purposes of the study to ensure the confidentiality and anonymity of the participants.

## Results

A total of 302 deceased relatives or caregivers were contacted and consented to conduct the VA interview, with three out of the respondents being female. Of these interviews, which were completed successfully over 4 months, InterVA-5 assigned either single, two, or three likely causes of death; a single cause of death to 299 individuals (99%), two likely causes to 26 individuals (8.60%), and three likely causes to 2 cases (0.66%). Three cases (0.99%) were explicitly designated ‘indeterminate’ by InterVA-5, generally reflecting a lack of particular VA data or contradictory evidence.

For the VA interviews where FRCoD was reported, 255 families (84.43%) gave a single cause of death, and 124 families (41.05%) reported two causes. There were 36 families (11.92%) where the given likely causes of death were considered ‘indeterminate’ since they did not match with the International Classification of Diseases (ICD-10) codes.

The results from a multivariable logistic regression model for exploring factors of determining the community perception are presented in Table 1. Based on the study findings, the odds ratio of the agreement decreases if the deceased was male compared to female (aOR: 0.52; 95% CI: 0.26 –1.03) and it significantly increases if married compared to single (aOR: 2.13; 95% CI: 1.02 –4.42). Improved perception of causes of death was associated with deaths of people with advanced education compared to illiterate (aOR:1.21; 95% CI: 0.45 – 3.20), and the death occurring at hospital compared to deaths at home (aOR: 1.25; 95% CI: 0.56 – 2.80), despite being not statistically significant. In addition, the odds of agreement tend to decrease if the interview was conducted with a respondent of a second-degree relationship to the deceased compared to a first-degree relationship (aOR: 0.91; 95% CI: 0.37 –2.20). Our results found that the odd ratio of the agreement for age groups (60-69, 70–79, and >=80 years) increases if the deceased aged 60-69 and 70-79 years compared to deaths aged 34-59 years, and the likelihood tends to decrease if the deceased was >=80 years compared to the reference (aOR: 1.34; 95% CI: 0.60 – 2.98, aOR:1.04; 95% CI: 0.45 – 2.41 and, aOR:0.48; 95% CI: 0.16 – 1.38, respectively).

**Table 1.** Background and characteristics of 302 deaths in Makkah region, Saudi Arabia, and showing multivariable OR for agreement between FRCoD and VACoD.

| Background characteristics | Number of deaths<br>*n* (%) | Crude OR (95% CI) | Adjusted OR (95% CI) |
| --- | --- | --- | --- |
| **Age group** |  |  |  |
| 34–59 | 68 (22.52) | Ref | Ref.<sup>a</sup> |
| 60-69 | 88 (29.14) | 1.37 (0.61 - 3.04) | 1.34 (0.60-2.98) |
| 70–79 | 84 (27.81) | 1.09 (0.48 – 2.51) | 1.04 (0.45-2.41) |
| >=80 | 62 (20.53) | 0.50 (0.17 - 2.42) | 0.48 (0.16-1.38) |
| **Sex** |  |  |  |
| Female | 126 (41.72) | Ref | Ref.<sup>b</sup> |
| Male | 176 (58.28) | 0.72 (0.40 - 1.31) | 0.52 (0.26-1.03) |
| **Respondent** |  |  |  |
| First degree relationship | 260 (86.09) | Ref | Ref.<sup>c</sup> |
| Second degree relationship | 42 (13.91) | 0.90 (0.37 - 2.16) | 0.91(0.37-2.20) |
| **Education** |  |  |  |
| Illiterate | 121 (40.07) | Ref | Ref.<sup>d</sup> |
| Basic education | 137 (45.36) | 1.09 (0. 51 - 2.32) | 1.09 (0.52- 2.30) |
| Advance education | 44 (14.57) | 1.24 (0. 46 - 3.32) | 1.21 (0.45- 3.20) |
| **Year of death** |  |  |  |
| 2018 | 28 (9.27) | Ref | Ref.<sup>c</sup> |
| 2019 | 69 (22.85) | 0.48 (0.14 - 1.54) | 0.49 (0.15-1.62) |
| 2020 | 161 (53.31) | 0.90 (0.34 - 2.42) | 0.90 (0.33-2.47) |
| 2021 | 44 (14.57) | 0.81 (0.24 – 2.66) | 0.89 (0.26-2.99) |
| **Marital status** |  |  |  |
| Single | 116 (38.41) | Ref | Ref. <sup>c</sup> |
| Married | 186 (61.59) | 1.78 (0.93 – 3.41) | 2.13 (1.02-4.42)* |
| **Place of death** |  |  |  |
| Home | 60 (19.87) | Ref | Ref. <sup>c</sup> |
| Hospital | 242 (80.13) | 1.29 (0.59 - 2.82) | 1.25 (0.56-2.80) |

**Abbreviation(s):** FRCoD: Family reported causes of death
VACoD: Verbal autopsy causes of death
a, adjusted for sex
b, adjusted for education, marital status and age groups
c, adjusted for age groups and sex
d, adjusted for age groups, sex and marital status

To evaluate the community perceptions of causes of death, CSMFs obtained from the VA using the InterVA-5 model were utilized to assess FRCoD, along with the CSMF absolute differences and their 95% confidence intervals. Table 2 shows that infectious diseases, stroke, and circulatory-related deaths were found to be underestimated as probable causes of death reported by families in relation to the VA reference standard. In contrast, CSMF of indeterminate and renal-related deaths groups, were overestimated as probable causes of death. In addition, accidental, NCD and diabetes related deaths groups were slightly overestimated with a minimal absolute CSMF difference. Despite no ICD codes being assigned by InterVA-5 for covid-19 (had no defined ICD code at the time) and mental illness-related deaths categories, the CSMF attributed to covid-19 by families was as high as 13.18% of all deaths.

**Table 2.** Cause specific mortality fractions per disease categories derived from VACoD and FRCoD for 302 deaths, Makkah region, Saudi Arabia.

| Categories | VACoD | FRCoD | Absolute Difference<br>95%CI |
| --- | --- | --- | --- |
| Infectious | 10.2 | 2.0 | -8.2 (-12.0,-4.4)* |
| Cancer | 5.2 | 4.4 | -0.8 (-4.2,2.6) |
| Diabetes | 14.3 | 15.8 | 1.5 (-4.2,7.2) |
| Circulatory | 35.7 | 19.7 | -16.0 (-23.0,-9.0)* |
| NCD | 4.5 | 4.9 | 0.4 (-3.0,3.8) |
| Renal | 2.8 | 9.2 | 6.4 (2.6,10.2)* |
| Accidental | 2.5 | 1.0 | -1.5 (-3.6,0.6) |
| Indeterminate | 7.7 | 16.8 | 9.1 (3.9,14.3)* |
| Stroke | 16.6 | 9.4 | -7.2 (-12.5,-1.9)* |
| Covid-19 | 0.0 | 13.1 | 13.2 (9.4,17.0)* |
| Mental | 0.0 | 3.1 | 3.1 (1.1,5.1)* |

**Abbreviation(s):** FRCoD: Family reported causes of death
VACoD: Verbal autopsy causes of death
\*Significant Absolute Difference

Based on findings from the CCC, there was an overall agreement between the two approaches (VACoD and FRCoD) in terms of cause-specific mortality. The Concordance of FRCoD concerning VACoD and fractions is shown in (Fig 1), with a CCC of 0.60 (95% CI: 0.20-1.00; p=003).

**Fig 1.**
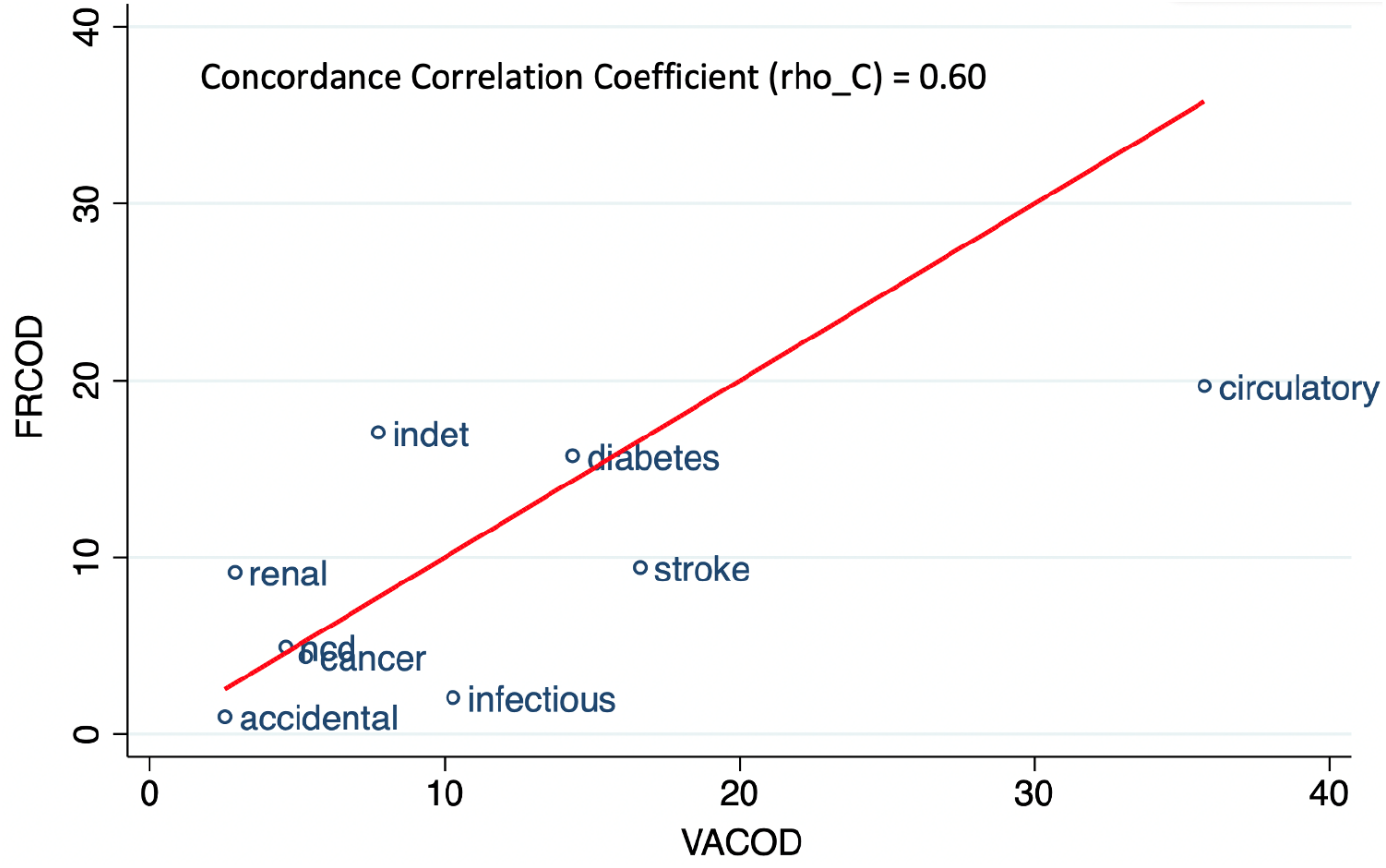
Concordance correlation coefficient (log-logscale) between CSMF determined by VA and respondent reports, in relation to the line of equivalence, for 302 deaths in Makkah city, Saudi Arabia.

## Discussion

The year 2030 is expected to be an exceptional milestone for Saudi Arabia, with a comprehensive agenda involving the assessment of essential national and global goals. The 17 Sustainable Development Goals (SDGs) including goal target 3.7 (*ensuring universal access to health-care services including for family planning, information, and education as well as vaccination*), and goal target 3.8 (*achieving Universal Health Coverage*), as well as the Saudi National 2030 Vision goals will both be assessed and disseminated. Creating partnerships with the private sector, academia, and civil society with an emphasis on strengthening civil registration and vital statistic systems (CRVS) would be crucial elements for succeeding in these national and global goals. However, gaps in registering vital data persist (a main task attributed to the CRVS), and despite increasing calls for employing standardized VA concepts to strengthen public health applications, including the CRVS, the VA approach is entirely novel in Saudi Arabia and many other neighboring countries where gaps in causes of death information exist. Exploring the perception of public health aspects, essentially how causes of death are perceived within communities in Saudi Arabia is crucial while the country’s health system is now undergoing a significant re-engineering process towards national and global agenda.

This study findings show a generally broad spectrum of community perceived mortalities with some crucial misconception based on the type of death and the background and characteristics of the deceased. During the data collection period, the world has experienced a devastating COVID-19 pandemic, which has ultimately re-profiled deaths and influenced the public and medical opinions of causes of death in Saudi Arabia and elsewhere. While the concept of community perception assessment is not new, understanding how local communities perceive cause of deaths, and consequently recognizing their associated risk factors is now timely and more crucial in view of the health and health system transitions due to COVID-19. Proposing such a standardized assessment would also be of interest to the Saudi health system to continuously monitor the health literacy of the people particularly within such dynamic Saudi communities. Based on published reports, low health literacy – which is how people are able to access, understand and utilize information and facilities to guide health-related decisions for themselves and other members in their communities – contributes to a wide range of health problems and is associated with poor health outcomes and ineffective chronic diseases management and treatments, such as the case of diabetes [16, 42, 43].

The study utilized VA to explore the community perceptions, which is mostly amenable for estimating CSMFs at population level and can adequately direct intended funds and interventional public health programs [44-46]. This study findings showed relatively minimal community misconception based on the derived CSMFs across diabetes, NCD, and accidental causes of death categories, implying that communities were more aware of and able to plausibly recognize these diseases as ultimate causes of death. Diabetes as a chronic illness demands frequent presentation at the healthcare services for routine follow up and treatments. High level of exposure to valid health information via professional medical practitioners can positively contribute to the community health literacy level, which may explain the higher level of perception for this cause of death category and likely other diseases from the NCD category. It can be said that communities are influenced by what they remember of signs and circumstances of death events whereas other factors such as exposure to local beliefs or to professional health practices can influence the subject’s memory [47]. For a disease such as cancer for instance, mental representations of death can be exceptionally influential and highlight how communities may perceive some inevitable illnesses [48]. This study shows trivial CSMF differences due to cancer, where communities are arguably attributing cancer illness to death in a plausible manner [48, 49]. Features of illness representations involve views about a disease’s etiology, time course, and curability [48]. For example, if individuals link cancer with death, they may avoid seeking health care services. In fact, there is considerable evidence that a fatalistic view of cancer hinders adherence to screening guidelines [50] and imped engagement in cancer protective behaviours such as physical activity, healthy diet and avoid smoking [51, 52]. Hence, addressing fatalistic beliefs through communication about certain diseases like cancer can play an important role in cancer prevention measures. Stroke as a cause of death, was however underestimated by communities, a pattern found to be consistent with previous Saudi and non-Saudi studies [53-55]. Difficulties in recalling or recognizing disease symptoms, rather than a mere lack of knowledge, can possibly explain the observed pattern by communities in relation to stroke, but may also reflect a need for a more effective stroke promotional campaign.

Public awareness of diseases that are associated with diabetes mellitus, like renal (kidney) and circulatory diseases are important for public health interventional plans. Our results show that respondents tend to overestimate renal diseases as causes of death, and this pattern is consistent with prior cross-sectional research from Saudi Arabia [56, 57], which revealed a dearth of knowledge about renal diseases among the Saudi population. While the prevalence of renal diseases is relatively high in Saudi Arabia due to climatic, behavioral and genetic factors [58], the Saudi health system has a comprehensive treatment and follow up management, both in terms of dialysis services and kidney transplantation. This interaction between high disease prevalence and adequate healthcare and the fact that most of the study population are presumably diabetic (which is medically associated with renal diseases), seems to influence the community’s perception of causes of death related to renal diseases. Saudi communities have underestimated circulatory diseases’ cause of death as it was also observed in other published reports revealing low perception of cardiovascular causes of death [59-61]. This underestimation could be attributed to the disease complexity and general lack of information and awareness. Although this may seem plausible from a medical point of view, failing to recognize symptoms of common illnesses can have some public health implications since most deaths have taken place at hospital settings where relatives or caregivers are presumably informed about the illness of their deceased. This was also observed in other cross-sectional studies linking low health literacy skills with unrealistic risk underestimation [62, 63]. Another plausible explanation is that circulatory diseases are frequently asymptomatic with risk factors slowly progressing over time leading to delayed recognition and perception. In line with a previous study [64], which demonstrated a low awareness of major infectious diseases, the present study revealed a low level of awareness in relation to infectious diseases, which is what would be expected in such setting due to stigmatization, and discrimination. This pattern supports the findings of previous studies in Saudi that reported a statistically significant positive correlation between some infectious diseases and stigma [65-67]. Stigma and discrimination in communities present a risk when reporting causes of death based on region, sex, and ethnicity, especially deaths related to stigmatized diseases [68]. As such, stigma can hinder accurate public awareness of some diseases and act as a barrier to people adopting health-promoting behaviors, seeking health care, and adhering to treatment.

The study findings indicate that community perception tends to be low if the deceased was >=80 years compared to 34-59 years. Despite being insignificance, this may be due to older people being more likely to develop chronic conditions, leading to disease recognition complexity at this age. Consequently, relatives attribute death to “fate” and perceive it as a natural part of the aging process rather than attributing it to a specific cause. We also found that community perception improves significantly if the deceased were married compared to single. Possibly, this is because deceased married individuals have a larger support network, including their spouse, family, and friends. Despite the ethical implications, symptoms and medical history of the deceased may have been shared with relatives, enabling a more accurate determination of the cause of death. Nevertheless, it should be noted that the results may vary according to the particular population being studied and the research methods used.

Although VA is commonly conducted in a registered population (a population being routinely monitored over time and space such as the concept followed in health socio-demographic and surveillance system), our study utilized medical records from the Saudi regional diabetes register as a denominator and later applied the VA process. Thus, the study finding may be overly pessimistic and have caused overlapping across the death categories. The method’s accuracy of cause of death determination is highly dependent on the type of death, the quality of the interview, and the procedures used to assign causes of death [26, 69]. A variety of factors related to the interviewer, the respondent’s background, or both can impact the interview quality and thereafter the VA results. The method has been shown to be reasonably practical for identifying causes of death in infancy or due to specific conditions such as injuries or maternal causes. On the other hand, medical causes of adult deaths share symptom complexes, making it difficult to differentiate between various causes of death from such descriptions when based solely on symptoms provided by relatives or occasionally by non-relative such as employers or work colleagues mainly in deaths among expats. Subjects with type 2 diabetes mellitus, for example, have a two- to four-fold increased risk of myocardial infarction and sudden death compared to individuals without diabetes, whereby roughly half of all diabetes deaths co-exist with some types of cardiovascular diseases [70]. This demonstrates the disease’s complexities and how the community may fail to recognize the true causes of death without effective health promotion and educational programs.

Despite the use of an internationally recognized standard VA method, interpreting and generalizing such findings in view of the small sample size can be challenging. The VA telephone interviews were used instead of the traditional face-to-face method to comply with the imposed COVID-19 pandemic’s physical distancing measures at the time of the data collection. Nevertheless, numerous studies have found that telephone interviews are as effective as face-to-face interviews [71, 72]. According to one study, the results of VA interviews via phone calls were consistent with previous literature, which found that the telephone interview method to be feasible, accepted by caregivers and healthcare workers, and has a reliable level of data quality [73]. Given the ethical nature of VA, it can arguably be said that conducting distant VA interviews on a fairly small sample in an area where VA has never been practiced before can be viewed as a strength in this study from an ethical perspective, which has the potential of reducing unwanted emotional or cultural consequences.

## Conclusion

Diabetes Mellitus is a leading cause of death globally. The Saudi community perception of causes of death was relatively plausible but varied substantially based on the type of death, sex, age >=80 years and other vital events like marital status. Higher or lower community perception is a proxy of how people may perceive risk factors associated with the causes of death, which can guide public health interventional programs. Unlike other inconsistent cross-sectional studies, this study proposed a standardized and sustainable method for monitoring people’s health literacy and allows harmonized comparison over time and space in otherwise unexplored population. While this study into VA respondents’ perceptions of causes of death is interesting for diabetes-related deaths, communities generated a considerably less complete and consistent picture of other cause-specific mortality. This highlights the importance of integrating rigorous and consistent VA methods within the routine medical services rather than relying on individual opinions mainly for complex causes of death outcomes. Nevertheless, translating cause-specific mortality data into effective policymaking remains challenging without standardized assessment of non-medical causes such as social and health system aspects contributing to death.

## Data Availability

The Study data are shared for reviewing purposes only.

https://github.com/FalehSaud/VA-data-.git

## Acknowledgments

The authors gratefully acknowledge all the deceased’s relatives who consented to involvement in this study. We also appreciate the Saudi Ministry of Health, Makkah Health Affairs, for approving this study.

## Supporting information

**S1 Table.**
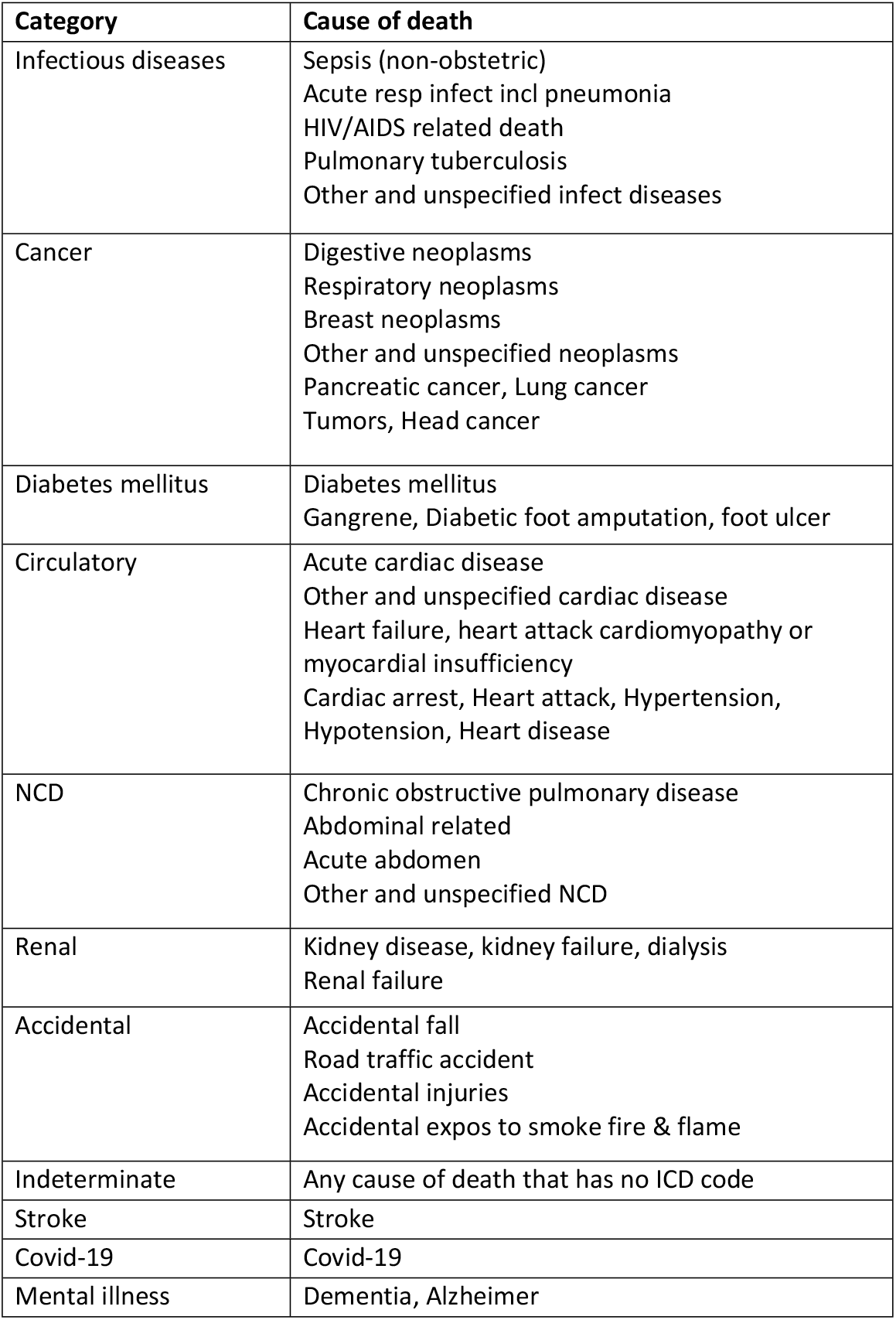
Categorizing of causes of death reported by VA and families.

